# Prevalence of Self Reported Auditory Dysfunction Among Call Center Employees and Its Association with Shift Work Across Various Call Centers in Cairo, Egypt, March 2026

**DOI:** 10.64898/2026.11.12.26367860

**Authors:** Mohamed Hisham Hakim Hussein, Amira Abdel Karim, Aseel Hisham Halim

## Abstract

**Background:** The rapid, unregulated expansion of the call center industry in developing countries has increased occupational health risks for young adults, particularly with regard to auditory health.

Prolonged exposure to acoustic trauma, long working hours, and the high intensity nature of call center work can negatively affect young adults at the start of their working lives and may contribute to permanent noise induced hearing loss (NIHL). This damage is often irreversible but preventable if identified at an early stage. This study aims to evaluate the burden of auditory impairment among call center employees in Egypt and to propose evidence based mitigation strategies.

**Methods:** A pilot cross sectional study was conducted among employees of various medical call centers over approximately three months (January to March 2026). Participants were recruited through voluntary (self selected) sampling. Data were collected using a structured questionnaire on work history, and statistical analysis was performed to examine associations with work duration and intensity. Three semi structured, open ended interviews were conducted in March 2026 to explore the symptoms in greater depth.

**Results:** Forty six call center employees completed the questionnaire. The most frequently reported symptoms were mishearing words (82.6%), difficulty understanding speech in background noise (80.4%), needing others to repeat themselves (78.3%), and sensitivity to loud sounds (78.3%); 60.9% reported tinnitus.

Higher auditory symptom severity was associated with a history of acoustic shock (Mann Whitney U = 344.0, p = .004) and with greater cognitive symptom burden (Spearman’s ρ = .416, p = .004), but not significantly with shift type (p = .177), years of work (p = .885), or daily and weekly working hours. Qualitative interviews described auditory strain that builds through the shift, disrupted sleep, informal coping, and the absence of employer guidance or symptom reporting pathways.

**Conclusion and Recommendations:** Self reported auditory symptoms were common in this preliminary sample of call center employees and were associated with acoustic shock exposure and cognitive symptom burden, but not significantly with shift pattern or length of service. Because the study was cross sectional, relied on self reports, and used a small volunteer sample (pilot study), the findings cannot establish causation or true prevalence. Recommended measures include acoustic shock limiters with incident reporting, audiometric screening that prioritizes at risk employees, safe volume education with regular breaks, and educational programs on auditory hazards and early warning signs to prevent actual auditory damage. Larger multi site studies with audiometric confirmation are also needed.

## Introduction

Noise is defined as unwanted and disturbing sound (Occupational Safety and Health Administration, 2002). It is estimated that approximately 16% of adult hearing loss worldwide is attributable to occupational noise exposure (Nelson et al., 2005). Although occupational hearing loss is traditionally associated with the mining and manufacturing industries, the digital transformation of the global economy has created new high risk groups, most notably call center operators handling various accounts. Regardless of the industry, the resulting damage is the same: progressive hearing loss.

In developing countries, economic instability and limited employment opportunities push young adults and recent graduates toward careers in the telecommunications sector, particularly call center services. These roles are accessible because of minimal entry requirements and competitive wages compared with other service sectors, which makes them attractive short term employment options for young people.

Call center employees frequently work long shifts with rotating schedules, which may disrupt circadian rhythm and impair melatonin secretion, causing chronic fatigue and elevated blood pressure (James et al., 2017), along with a high incidence of psychological distress, anxiety, and burnout. Fatigue, sleep impairment, and disrupted melatonin secretion contribute to long term health deterioration (Toker & Güler, 2022).

From an auditory perspective, continuous exposure to unregulated noise from calls, call disconnections, and unexpected high intensity acoustic signals are stressors that may increase the risk of noise induced hearing loss (NIHL), tinnitus, and central auditory processing dysfunction.

### The Research Gap

Despite the rapid expansion of the call center industry, particularly with Egypt emerging as a regional hub for business process outsourcing and customer service operations, research on noise induced hearing loss among call center operators remains limited, particularly in the Middle East and Egypt. Previous studies have addressed related concerns in different populations (Pawlaczyk Luszczynska et al., 2018). For example, a study of call center operators in a European setting found that most operators had normal hearing, although a meaningful minority showed early high frequency notching despite reporting auditory and visual symptoms. This highlights the importance of subjective symptom reporting, since symptoms may occur before measurable hearing loss is detected, and the need for further studies. However, that study was conducted in a European context and did not specifically examine occupational hearing among call center workers in Egypt. Similarly, a recent review (Chen & Su, 2025) focused primarily on the mechanisms of auditory damage and noise induced hearing loss in broader workplace populations rather than specifically examining call center employees. Furthermore, limited attention has been given to the psychological and cognitive strain that may accompany auditory symptoms, and to early subjective indicators that may precede clinically detectable hearing impairment.

This gap directly shaped the objectives of the present study, which aimed to determine the prevalence and severity of self reported auditory symptoms among call center employees in Egypt and to identify the occupational and lifestyle factors that may be strongly associated with symptom severity. Rather than relying solely on audiometric assessment, the study adopted a symptom based approach to capture early auditory complaints and their relationship with cognitive and psychological symptom burden. The findings provide additional evidence from an understudied occupational population and suggest that auditory symptoms may be highly prevalent among call center employees. In the present sample, approximately 80% of participants reported mishearing and difficulty understanding speech in background noise, while a history of acoustic shock emerged as one of the strongest correlates of symptom severity. A significant association was also observed between auditory and cognitive symptom burden, suggesting that auditory difficulties among call center employees may extend beyond hearing related complaints alone.

### Study Objectives

The main objective of the study was to evaluate the extent of auditory dysfunction among call center operators in Egypt and to investigate the factors that may influence its prevalence and severity. The specific objectives were:

1. To identify the prevalence of self reported auditory dysfunction among call center employees, including tinnitus, difficulty hearing in background noise, and other early symptoms of hearing loss.
2. To assess the severity and pattern of hearing impairment, including which ear is more affected.
3. To examine the association between shift work and auditory dysfunction, focusing on whether night or rotating shifts increase the risk or severity.
4. To detect early warning signs and patterns of auditory and cognitive impairment, enabling early intervention and prevention strategies.
5. To explore lifestyle and occupational factors that contribute to auditory risk, including duration of headset use, personal audio device use, smoking, and past medical history.
6. To provide evidence based recommendations for mitigating noise related hearing damage in call center environments, including occupational health guidelines and employee awareness strategies.

## Methods

### Study Design

This is a pilot cross sectional study with a mixed methods component (a questionnaire survey and open ended interviews).

### Study Duration

January 2026 to September 2026 (including preparation, data collection, analysis, and reporting). Data collection took almost three months (January to March 2026), and the interviews were conducted in March 2026.

### Study Setting

The study was conducted among employees of various call center companies in the Greater Cairo area, Egypt, including Transcom, Elevate Call Center, and Concentrix Egypt.

Data were collected through online questionnaires and surveys.

### Study Population

Target population: call center employees in Egypt aged 18–40 years

Inclusion criteria:

- Current full time or part time call center employees
- Minimum of 6 months of employment
- Willingness to participate and provide informed consent

Exclusion criteria:

- Pre existing severe hearing loss unrelated to occupational exposure
- Recent ear surgery (<6 months)
- Chronic medical conditions that significantly affect hearing or equilibrium

### Sample Size and Sampling Method

#### Sample size

A target of 100–120 participants was set on the basis of feasibility. Reaching this target was not logistically feasible within the data collection period of almost three months, so the study is presented as a pilot study to inform a larger future study.

#### Recruitment

Participation was voluntary (volunteer, or self selected, sampling). The questionnaire was shared via WhatsApp with call center employees, and every employee who chose to respond was included. Many of the employees who were reached did not complete the questionnaire, and only completed questionnaires (n = 46) were analyzed.

#### Response and missing data

The exact number of employees who received the questionnaire link was not recorded, so a response rate could not be calculated. Incomplete questionnaires were excluded, and analyses of the completed questionnaires used available cases.

### Data Collection

#### a) Questionnaire

- A structured questionnaire (self administered or online) covering:

- Demographics (age, gender, education)
- Work history (duration of employment, work hours, shifts, headset use, loudness)
- Lifestyle factors (sleep, smoking, headphone use outside work, and listening habits)
- Past medical and hearing history (middle ear disease, ear surgery, hereditary disorders, cholesterol levels, arterial hypertension, head trauma)
- Physical features (body weight, height, skin pigmentation)
- Auditory symptoms (severity/discomfort scale, affected ear)
- Listening fatigue and cognitive load (severity/discomfort scale)
- Optional participation in follow up interviews
- Data handling: All responses were anonymized. Contact details for follow up were stored separately.

#### b) Open Ended Interviews (Qualitative Component)

- Purpose: To explore participants’ experiences in detail, focusing on:

- Perceived auditory strain and fatigue
- Sleep disturbance due to night shifts
- Coping strategies and headset use habits
- Any unreported or subtle symptoms not captured in the questionnaire
- Method:

- Three semi structured, open ended interviews were completed in March 2026 (originally planned as 3–5 focus group sessions of 5–8 participants each)
- Interviews followed a guide of 22 open ended questions adapted from questions used in published studies (Afifi et al., 2020; Jeon & Kim, 2022; Venet et al., 2018; see Appendix 2). The guide was reviewed by the study supervisor and by the Research Ethics Committee, Faculty of Medicine, Tanta University
- Duration: 30–45 minutes per interview
- Audio recorded and transcribed with participant consent
- The interviewer maintained participant confidentiality throughout
- Analysis:

- Thematic analysis to identify common patterns and experiences
- Integration of qualitative findings with quantitative data to enrich interpretation

### Ethical Considerations

- Ethical approval was obtained from the Research Ethics Committee, Faculty of Medicine, Tanta University (approval code: 36264PR1310159/11/26; date of approval: 5 November 2026; see Appendix 1)
- Participation was voluntary, and informed consent was obtained before the questionnaire
- Anonymity and confidentiality were maintained
- Participants could withdraw at any time without consequences
- The questionnaire and methodology were reviewed by the Department of Public Health and Community Medicine, Tanta University

### Data Management

All data collected from the online questionnaires were first entered into Microsoft Excel for cleaning and organization, then exported to SPSS for analysis.

Reporting guidelines: The quantitative component was reported with reference to the STROBE statement for cross sectional studies, and the qualitative component with reference to the COREQ checklist.

### Quantitative Data

- Descriptive statistics were calculated to summarize the demographic characteristics, occupational exposure (e.g., work hours, shift type, headset use), and auditory symptoms. Measures included range, mean, standard deviation, frequencies, and percentages. Ninety five percent confidence intervals for proportions were calculated using the Wilson score method.
- Individual symptom items were dichotomized into present/absent for prevalence estimates. Composite auditory (seven items) and cognitive (four items) symptom severity scores were calculated as the sum of the item scores, and their distributions were assessed with the Kolmogorov–Smirnov and Shapiro–Wilk tests. Analyses used available cases (one participant had missing cognitive data).
- Because symptom items are ordinal and several subgroups were small and unbalanced (e.g., five day shift employees), nonparametric tests were used: Spearman’s rank correlation for associations between symptom severity and continuous or ordinal exposures, the Mann–Whitney U test for two group comparisons (e.g., acoustic shock yes/no), and the Kruskal–Wallis H test for comparisons across three or more groups (e.g., day, night, and rotating shift). Statistical significance was set at a two sided p < .05.

## 1. Results

Forty six call center employees participated in the study and completed the self report questionnaire on auditory symptoms, cognitive symptoms, and occupational/lifestyle exposures. One participant had missing data on the cognitive symptom measure, giving an analytic sample of n = 45 for analyses involving the cognitive composite score; all other variables were complete for the full sample of 46.

### 1.1 Sample Characteristics

A total of 46 completed questionnaires were included in the analysis, and incomplete questionnaires were excluded. Table 1 summarizes the descriptive characteristics of the study sample, including anthropometric measures and reported occupational exposure variables (years of work, daily and weekly working hours, and sleep duration).

**Table 1.** Descriptive statistics of demographic and occupational characteristics (N = 46).

| **Variable** | **N** | **Min** | **Max** | **Mean** | **SD** |
|---|---|---|---|---|---|
| Age group (category) | 46 | 1.00 | 2.00 | 1.61 | 0.49 |
| Years of work (category) | 46 | 1.00 | 3.00 | 1.63 | 0.61 |
| Work hours/day (category) | 46 | 1.00 | 5.00 | 3.04 | 0.79 |
| Work hours/week (category) | 46 | 1.00 | 5.00 | 3.52 | 0.98 |
| Weight (kg) | 46 | 55.00 | 95.00 | 71.30 | 11.42 |
| Height (m) | 46 | 1.55 | 1.85 | 1.74 | 0.10 |
| Sleep duration (category) | 46 | 1.00 | 4.00 | 2.07 | 0.77 |

### 1.2 Objective 1 — Prevalence of Self Reported Auditory Symptoms

Individual symptom items were dichotomized into present/absent indicators. Self reported auditory symptoms were common across the sample. Mishearing words was the most frequently reported symptom (82.6%), followed by difficulty understanding speech against background noise (80.4%), needing conversations repeated (78.3%), and sensitivity to loud sounds (78.3%). Tinnitus was reported by 60.9% of employees, and a similar proportion (60.9%) reported increasing the volume of the television or personal devices beyond what others considered comfortable. Difficulty localizing the source of a sound was reported by 63.0% of the sample (Table 2, Figure 1).

**Figure 1.**
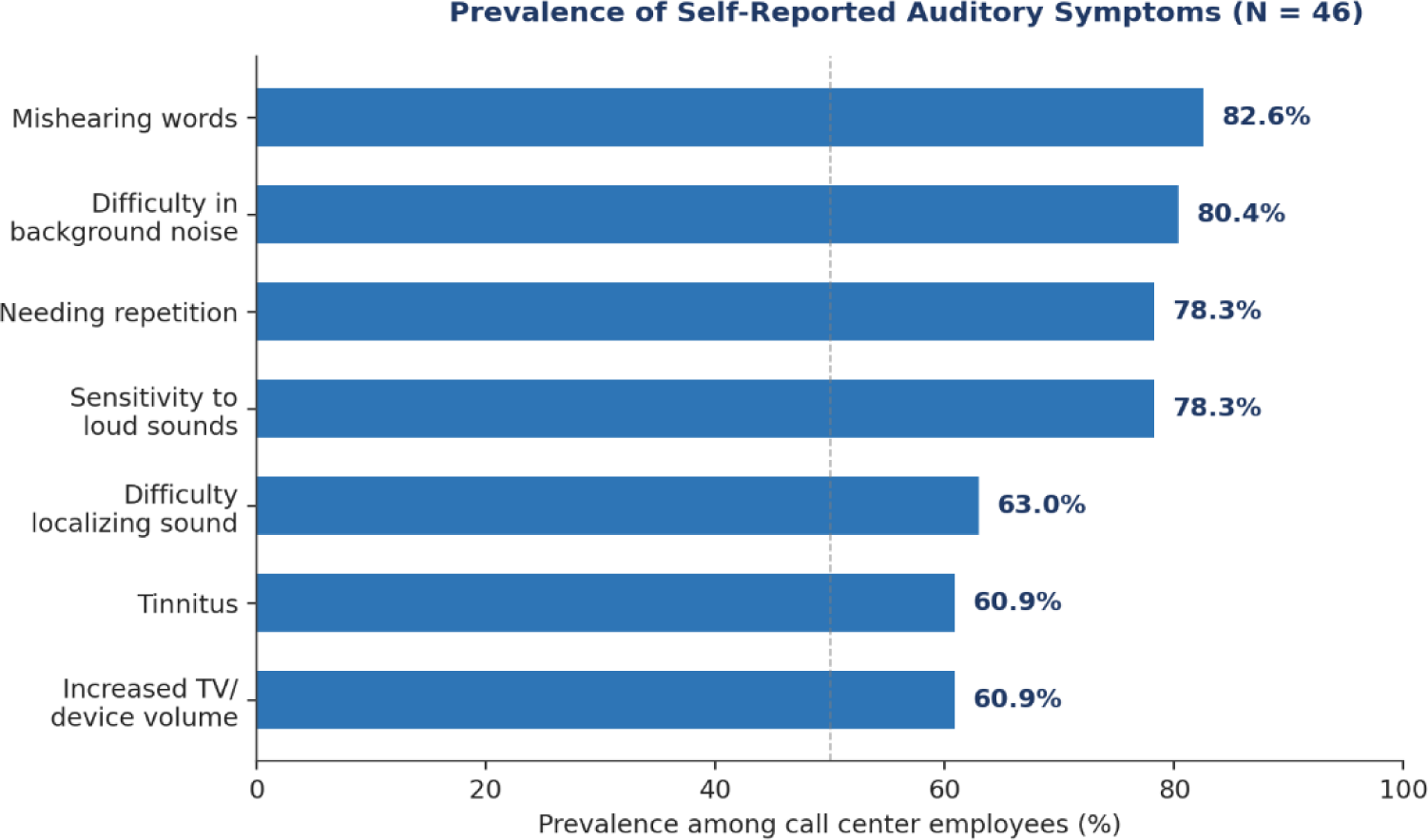
Prevalence of self reported auditory symptoms, ranked as the most to least common.

**Table 2.** Prevalence of self reported auditory symptoms among call center employees (N = 46).

| Symptom | Present, n (%) | Absent, n (%) |
|---|---|---|
| Mishearing words during conversation | 38 (82.6%) | 8 (17.4%) |
| Difficulty understanding speech with background noise | 37 (80.4%) | 9 (19.6%) |
| Needing others to repeat themselves | 36 (78.3%) | 10 (21.7%) |
| Sensitivity to loud sounds | 36 (78.3%) | 10 (21.7%) |
| Difficulty identifying where a sound is coming from | 29 (63.0%) | 17 (37.0%) |
| Tinnitus | 28 (60.9%) | 18 (39.1%) |
| Increased TV/device volume | 28 (60.9%) | 18 (39.1%) |

Because of the small sample (n = 46), these prevalence estimates are imprecise. The 95% confidence intervals (Wilson method) were: mishearing words 82.6% (95% CI 69.3–90.9); difficulty understanding speech in background noise 80.4% (95% CI 66.8–89.3); needing others to repeat themselves 78.3% (95% CI 64.4–87.7); sensitivity to loud sounds 78.3% (95% CI 64.4–87.7); difficulty identifying where a sound is coming from 63.0% (95% CI 48.6–75.5); tinnitus 60.9% (95% CI 46.5–73.6); increased television or device volume 60.9% (95% CI 46.5–73.6).

#### 1.2.1 Objective 1 (continued) — Prevalence of Self Reported Cognitive Symptoms

Individual symptom items were dichotomized into present/absent indicators. Self reported cognitive symptoms were common across the sample: headaches after prolonged headset use (86%), mental exhaustion after long listening tasks (86.3%), difficulty concentrating during long calls (83.3%), and slower processing of spoken information (68%) (Table 3, Figure 2).

**Figure 2.**
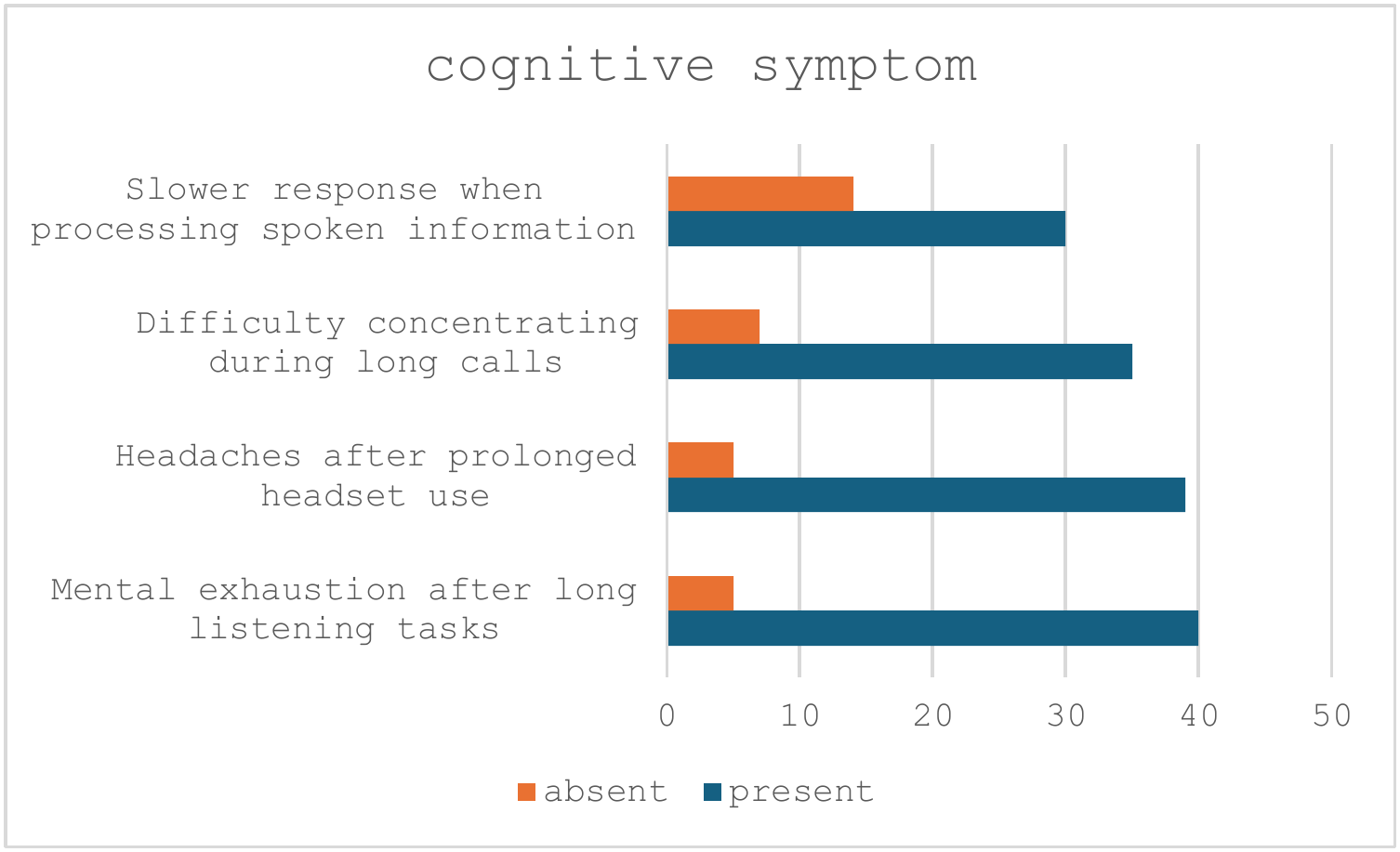
Prevalence of self reported cognitive symptoms among call center employees.

**Table 3.** Prevalence of self reported cognitive symptoms among call center employees.

| Symptom | Present, n (%) | Absent, n (%) |
|---|---|---|
| Mental exhaustion after long listening tasks | 40 (86%) | 6 (14%) |
| Headaches after prolonged headset use | 39 (88%) | 5 (12%) |
| Difficulty concentrating during long calls | 35 (83%) | 7 (17%) |
| Slower response when processing spoken information | 30 (68%) | 14 (32%) |

### 1.3 Objective 2 — Severity and Pattern of Auditory Impairment

A composite auditory symptom severity score (sum of the seven symptom items) was computed for each participant. The composite score approximated a normal distribution (Kolmogorov–Smirnov p = .113; Shapiro–Wilk p = .237), with a mean of 17.59 (SD = 6.20, 95% CI: 15.75–19.43), a median of 18.0, and scores ranging from 7 to 33 (Figure 3). The distribution was slightly right skewed (skewness = 0.194) with near normal kurtosis (−0.549), consistent with a moderate overall symptom burden and a smaller subgroup of employees reporting markedly higher severity.

**Figure 3.**
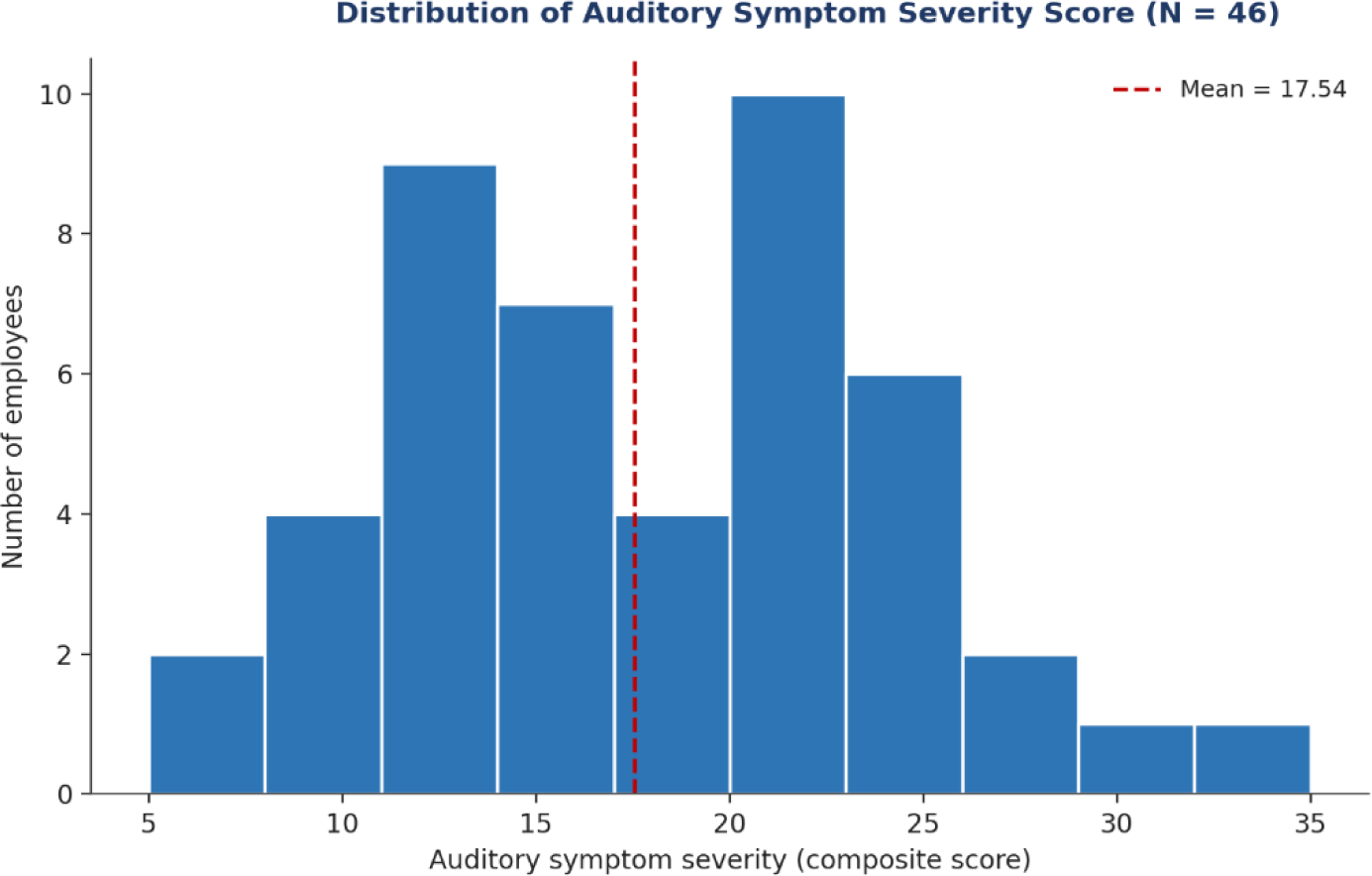
Distribution of composite cognitive symptom severity scores (n = 45).

Severity was also examined by affected ear. A boxplot comparison indicated differences in the spread of scores across the three ear categories; however, this apparent pattern is descriptive only and requires confirmation with a formal Kruskal–Wallis test before it can be reported as a definitive finding.

### 1.4 Objective 4 — Cognitive Symptoms and Their Relationship to Auditory Dysfunction

A parallel composite score for cognitive symptoms (mental exhaustion, headache, difficulty concentrating, and slowed processing) was calculated. This score was likewise approximately normally distributed (Shapiro–Wilk p = .428), with a mean of 12.11 (SD = 3.84, 95% CI: 10.96–13.27), median 13.0, and a range of 4 to 20 (n = 45) (Figure 4).

**Figure 4.**
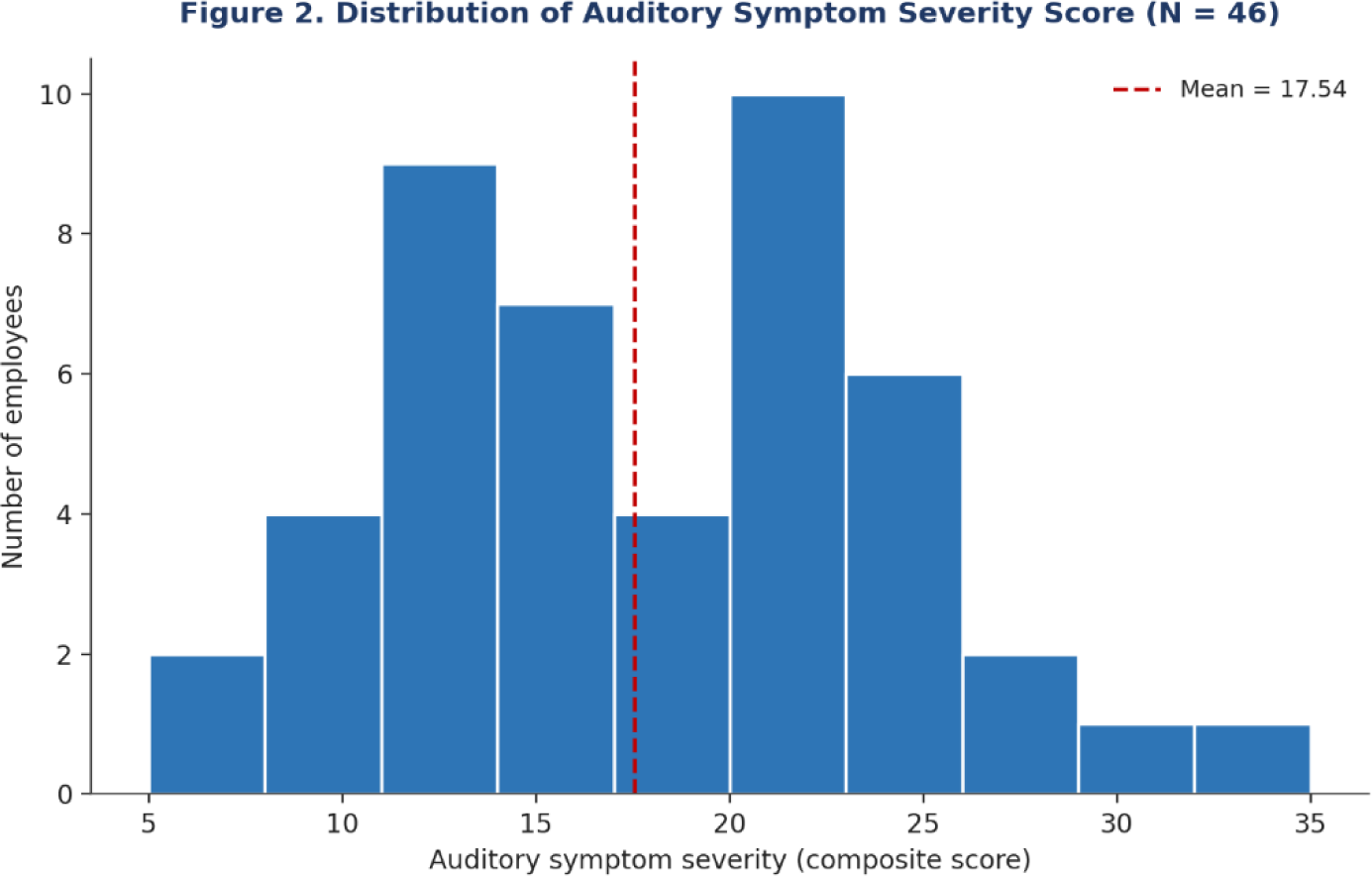
Distribution of composite auditory symptom severity scores (N = 46).

**Figure 5.**
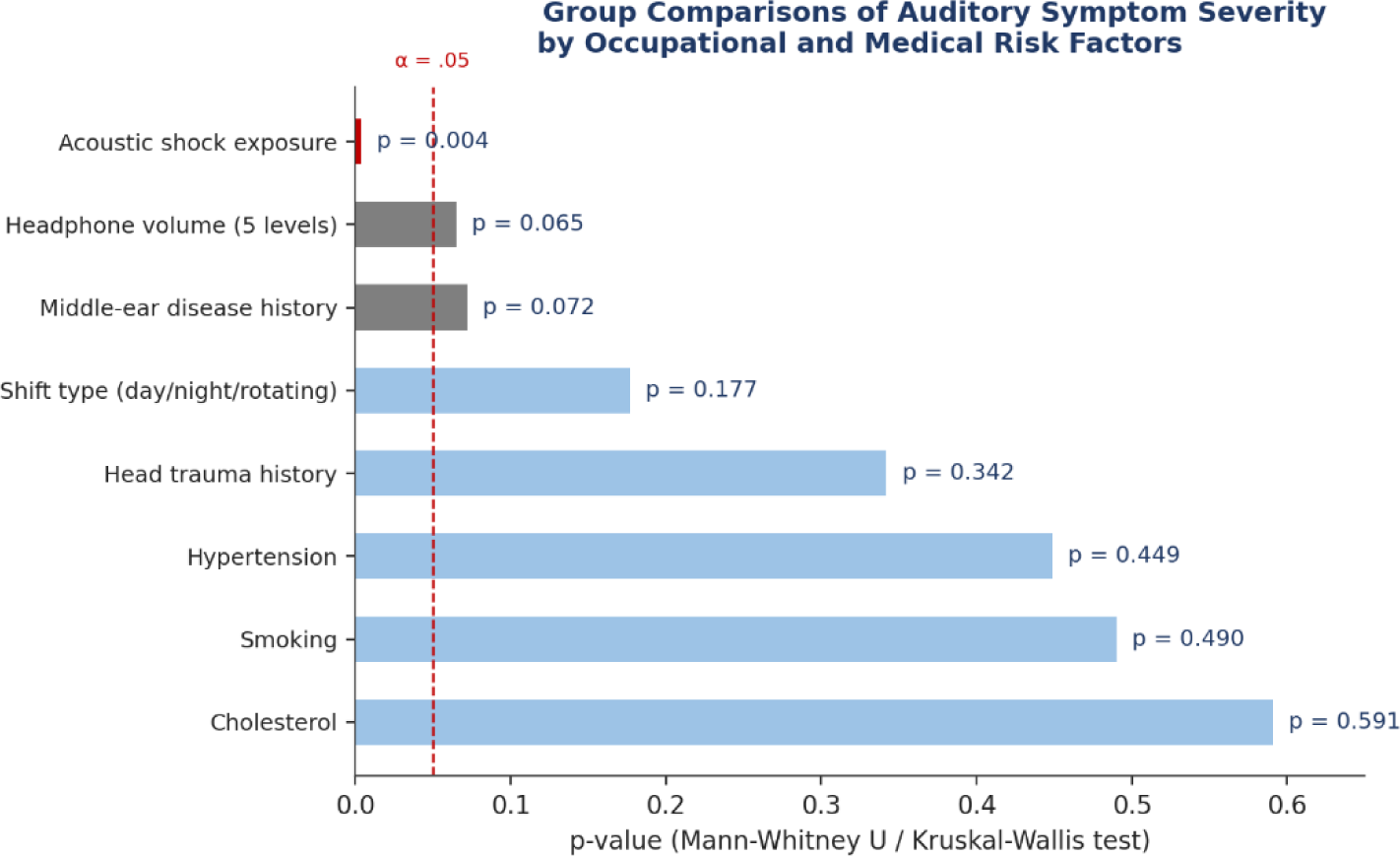
Summary of group comparison test results (Mann–Whitney U / Kruskal–Wallis) for auditory symptom severity across occupational and medical risk factors. Dashed red line marks the α = .05 significance threshold.

A Spearman correlation showed a significant, moderate positive association between auditory and cognitive symptom severity (ρ = .416, p = .004, n = 45), indicating that employees who reported more severe auditory symptoms also tended to report greater cognitive strain. Examining the individual cognitive items, slowed information processing showed the strongest association with auditory severity (ρ = .500, p < .001), while difficulty concentrating (ρ = .287, p = .053) and headache (ρ = .282, p = .061) showed borderline, non significant trends, and mental exhaustion was not significantly associated (ρ = .213, p = .154). The cognitive items also correlated with one another (ρ = .417–.587, p ≤ .004), suggesting they reflect a coherent underlying construct of listening related cognitive load.

### 1.5 Objective 3 — Shift Work and Auditory Dysfunction

Composite auditory symptom severity did not differ significantly across shift type (day, night, or rotating shift) (Kruskal–Wallis H(2) = 3.461, p = .177). This finding does not support shift timing, in itself, as a determinant of auditory symptom severity in this sample; cumulative exposure factors examined below (e.g., acoustic shock history) appeared more strongly related to symptom severity than shift schedule.

#### 1.5.1 Cognitive Symptoms by Shift Type

The distribution of employees across shift categories was unbalanced: 5 employees (10.9%) worked day shift, 12 (26.1%) worked night shift, and 29 (63.0%) worked a rotating shift (Figure 6).

**Figure 6.**
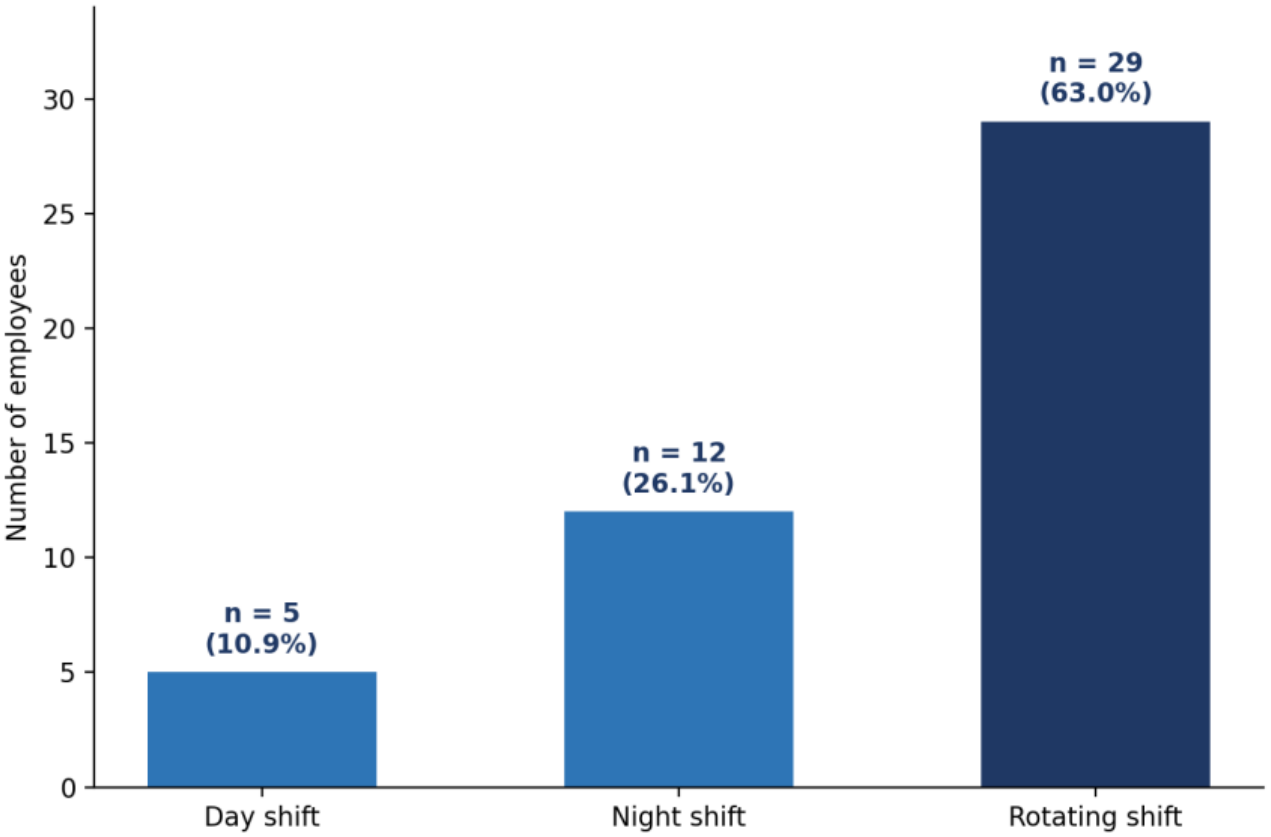
Distribution of employees by shift type (N = 46).

**Figure 7.**
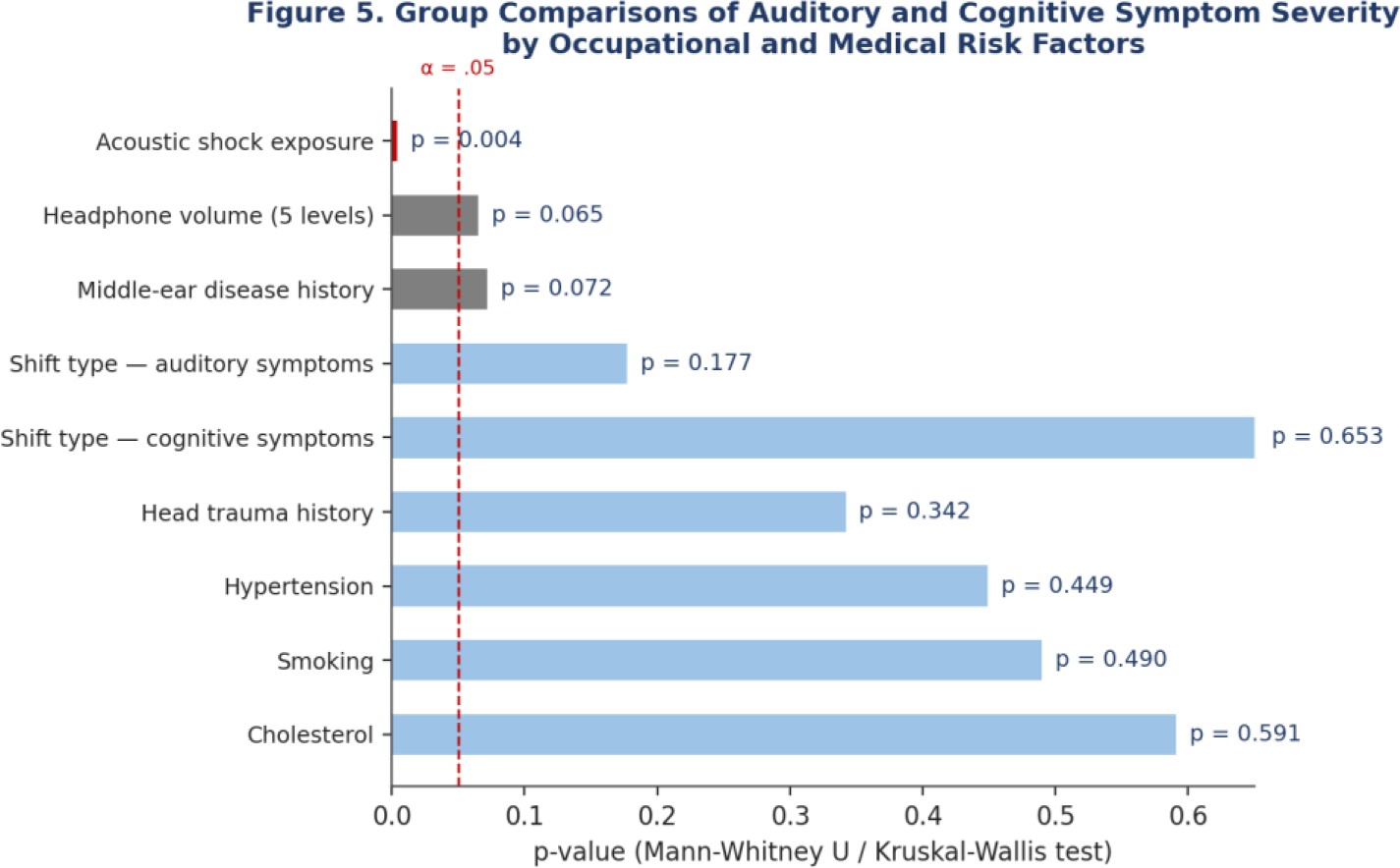
Summary of group comparison test results (Mann–Whitney U / Kruskal–Wallis) for auditory and cognitive symptom severity across occupational and medical risk factors. Dashed red line marks the α = .05 significance threshold.

Cognitive symptom severity was compared across the three shift categories using the Kruskal–Wallis H test, the appropriate nonparametric test for a grouping variable with more than two levels. The test showed no significant difference in cognitive symptom severity across shift categories (H(2) = 0.851, p = .653, n = 45), and the null hypothesis was retained.

This mirrors the finding for auditory symptom severity by shift (Section 1.5, p = .177): neither auditory nor cognitive symptom burden differed meaningfully by shift schedule in this sample. Taken together, shift timing does not appear to be a driving factor behind either symptom domain, reinforcing that acoustic shock exposure and the auditory–cognitive symptom link (Sections 1.4 and 1.6) are more informative targets for prevention than shift pattern. As with the auditory comparison, the small and unbalanced subgroup sizes (day shift, n = 5) limit the power of this test to detect a true difference, so this should be interpreted as an absence of evidence for a shift effect rather than confirmed evidence of no effect.

### 1.6 Objective 5 — Lifestyle and Occupational Risk Factors

Associations between auditory symptom severity and a range of occupational, lifestyle, and medical history variables were tested using Spearman correlation (continuous/ordinal exposures) and Mann–Whitney U or Kruskal–Wallis tests (categorical exposures). Results are summarized in Table 4, Table 5, and Figure 8.

**Figure 8.**
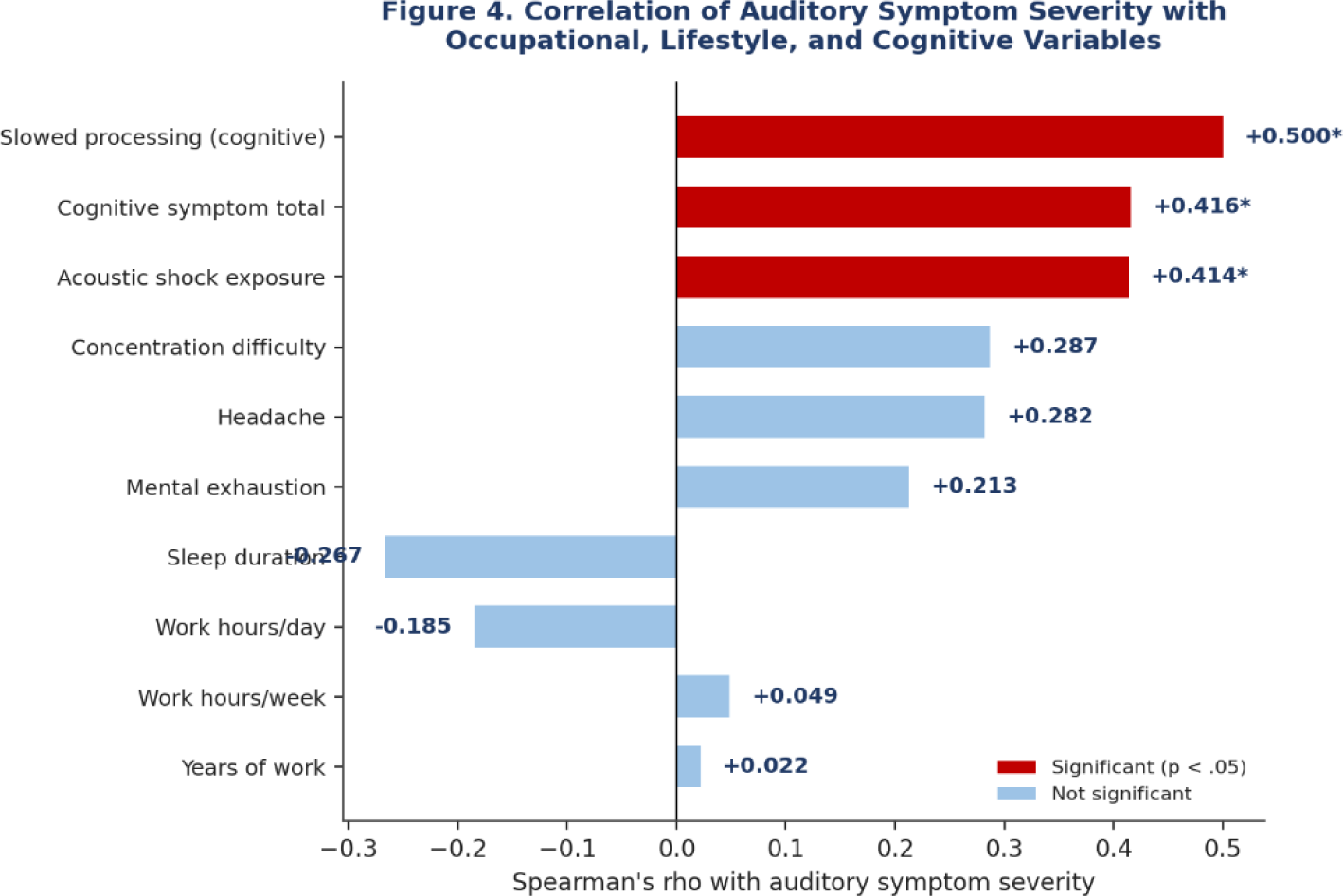
Spearman correlations between auditory symptom severity and occupational, lifestyle, and cognitive variables. Red bars denote statistically significant correlations (p < .05); asterisk marks p < .05.

**Table 4.** Spearman correlations between auditory symptom severity and occupational/lifestyle exposures. ** p < .01.

| Variable | Spearman's ρ | p value | N |
|---|---|---|---|
| Acoustic shock exposure | 0.414 | .004 ** | 46 |
| Sleep duration | −0.267 | .073 | 46 |
| Work hours/day | −0.185 | .217 | 46 |
| Work hours/week | 0.049 | .744 | 46 |
| Years of work | 0.022 | .885 | 46 |

**Table 5.** Group comparisons of auditory symptom severity by categorical risk factors. ** p < .01.

| **Variable** | **Test** | **Statistic** | **p value** |
|---|---|---|---|
| Acoustic shock exposure (yes/no) | Mann–Whitney U | U = 344.0 | .004 ** |
| Headphone/device volume (5 levels) | Kruskal–Wallis | H(4) = 8.833 | .065 |
| Middle ear disease history | Mann–Whitney U | U = 248.0 | .072 |
| Shift type (3 levels) | Kruskal–Wallis | H(2) = 3.461 | .177 |
| Head trauma history | Mann–Whitney U | U = 107.0 | .342 |
| Hypertension | Mann–Whitney U | U = 124.5 | .449 |
| Smoking status | Mann–Whitney U | U = 180.5 | .490 |
| Cholesterol history | Mann–Whitney U | U = 54.5 | .591 |
| Ototoxic medication use | Mann–Whitney U | not computable (single category) | — |

The clearest risk factor to emerge was a history of acoustic shock exposure (e.g., a sudden loud sound through the headset): employees who reported such an event had markedly higher auditory symptom severity than those who did not (mean rank 27.25 vs. 14.93; Mann–Whitney U = 344.0, standardized test statistic = 2.87, p = .004), and this was corroborated by a significant correlation (ρ = .414, p = .004). Headphone/device volume level showed a borderline association that did not reach conventional significance (p = .065), and a history of middle ear disease showed a similar borderline trend (p = .072, exact test), with affected employees showing a higher mean rank (30.30 vs. 21.61) — both worth flagging as trends given the modest subgroup sizes. Sleep duration showed a weak, non significant inverse trend (ρ = −.267, p = .073), suggesting a possible but unconfirmed link between shorter sleep and greater symptom severity. No significant associations were found for years of work, daily or weekly work hours, smoking status, cholesterol, hypertension, or head trauma history (all p > .05); it should be noted that several of these subgroups (cholesterol, n = 2; hypertension, n = 5; head trauma, n = 4) were very small, limiting statistical power. Ototoxic medication exposure could not be tested because all participants fell into a single category.

### 1.7 Objective 6 — Implications for Prevention

Taken together, the results point to acute acoustic events (acoustic shock) and cumulative listening load (reflected in the auditory–cognitive symptom correlation) as the two most actionable targets for intervention, rather than shift schedule or duration of employment per se. These findings are developed further in the Discussion and used to frame the evidence based recommendations in Section 2.6.

#### Thematic Analysis of Interview Data: Occupational Auditory Strain, Sleep Disruption, and Psychological Load Among Call Center Workers

##### Method Note

Three semi structured, open ended interviews (Participants 1, 2, and 3) were conducted in March 2026 using an interview guide adapted from questions used in the published literature (Appendix 2). The guide covered four a priori topic areas: (1) perceived auditory strain and fatigue, (2) sleep disturbance due to night shifts, (3) anxiety, mental fatigue, coping strategies, and headset use habits, and (4) unreported or subtle symptoms. Transcripts were bilingual (Arabic–English) and machine transcribed. Two of the three transcripts (those of Participants 2 and 3) contained substantial automatic speech recognition (ASR) errors and unsegmented speech, so close paraphrase was used instead of direct quotation for those sections, consistent with standard practice for degraded qualitative data. Analysis followed Braun and Clarke’s (2006) six phase approach to thematic analysis: familiarization, initial coding, theme searching, theme review, theme definition and naming, and write up.

Participants: P1 — customer service/back office agent (fixed day shift, later flexible hours); P2 — call center agent (rotating shifts); P3 — sales agent (rotating/night shifts, high pressure targets).

### Theme 1: Auditory Strain Accumulates Progressively Across the Shift

All three participants described a clear within shift trajectory rather than a static level of discomfort — symptoms were mild or absent at shift start and intensified toward the end.

#### Sub theme 1.1 — Fatigue proportional to call intensity

P1 distinguished a high volume “frontline” role (constant, back to back calls) from a self paced back office role and reported that auditory and physical fatigue (“awful headaches,” needing painkillers) were markedly worse during the frontline period. P3 similarly linked ear pain directly to shift progression, describing mild discomfort early on that became a “slight pain” by the end of the shift, and explicitly confirmed that this was shift related rather than incidental.

#### Sub theme 1.2 — Compensatory volume escalation

Both P2 and P3 reported raising the headset volume as the shift progressed in order to keep understanding callers. P3 described pushing the volume to very high levels through a browser extension in the later hours of the shift, framing it as a direct response to reduced clarity rather than a stable preference.

#### Sub theme 1.3 — Tinnitus, pressure, and ear clogging sensations

P1 reported a progression from occasional muffled hearing to persistent ringing (tinnitus) that continued even after leaving the job, alongside a sensation of pressure or clogging localized to the ear kept under the headset. This was severe enough to prompt a medical visit, but the examination performed (visual otoscopy) did not include audiometric or frequency testing. This is a methodological gap, since no objective threshold data exist to corroborate the self report. P2 and P3 reported pressure sensations during or after calls but were less certain about buzzing specifically; they noted that symptoms correlated with, and increased with, shift intensity.

### Theme 2: Sleep Is Structurally Disrupted by Shift Demands, Independent of “Night Shift” Framing

Sleep disruption was reported across all three participants, but the mechanism differed by role rather than by a simple day/night distinction.

#### Sub theme 2.1 — Reduced sleep volume and duration under full time and rotating schedules

P1 reported that sleep was largely stable during a hybrid schedule but deteriorated once physical, symptom related discomfort (ear pressure) began to interfere, more than because of the shift timing itself. P3 reported a much sharper effect: sleep dropped from a baseline of about 9 hours to 5–6 hours of poor quality sleep under a full overnight schedule, and P3 described daytime recovery sleep as never equivalent to true rest. P2 described the lack of a clear sleep schedule, difficulty coping with shift work, and progressively decreasing sleep hours.

#### Sub theme 2.2 — Downstream effects on functioning

P3 linked poor sleep directly to reduced work performance in a role that “needs you to be…full of energy,” and to broader emotional overload and a reduced capacity to manage daily life outside work. P1 did not report the same degree of cognitive or mood impact from sleep specifically, attributing daytime tiredness more to cumulative workload than to sleep loss per se.

### Theme 3: Psychological Load Is Role Dependent, With Performance Linked Anxiety Concentrated in Sales

#### Sub theme 3.1 — Anxiety tied to income and performance stability

P3, who works in a target driven sales role, reported that anxiety was conditional on performance (“when I’m doing well at work I don’t feel that anxious”) and directly linked poor sleep and fatigue to fear over job and income stability. On the structured symptom checklist, P3 confirmed difficulty concentrating and trouble sleeping but denied most somatic anxiety symptoms (sweating, trembling, palpitations).

#### Sub theme 3.2 — Interference with life outside work

P3 described concrete downstream effects, including missed social plans and difficulty attending personal obligations, and attributed these explicitly to rotating and overnight shift patterns rather than to the sales role alone.

#### Sub theme 3.3 — Normalization among staff without performance targets

P1’s account (customer service, not commission or target based) framed the stress as diffuse and comparative (“stressful from all sides”: study, work, and life stage) rather than attributing it to a single dominant cause. P1 also noted that colleagues who treated the job as a primary income source seemed comparatively more at ease with the workload.

### Theme 4: Coping Is Informal, Substance Assisted, and Entirely Self Directed

#### Sub theme 4.1 — Self directed micro breaks

P1 described discretionary strategies, such as stepping away from the desk, eating, and taking short naps, used opportunistically rather than as part of any structured break policy.

#### Sub theme 4.2 — Stimulant reliance

P3 reported becoming a heavy smoker and relying on caffeine (espresso shots) specifically to manage shift induced exhaustion, framing this as a direct coping response to the energy demands of the sales role.

#### Sub theme 4.3 — Unilateral headset use as informal ear “resting”

Both P1 and P2 reported habitually keeping one ear free of the headset (typically the non dominant or left ear) to preserve situational awareness, switching sides when the free ear became uncomfortable. This suggests an unprompted, self taught compensatory behavior rather than anything trained or company issued.

#### Sub theme 4.4 — Absence of employer guidance

All participants independently confirmed that no formal guidance existed on headset volume limits or break frequency. P1 noted that the headset hardware had a built in, color coded loudness indicator, but no policy enforced attention to it. P3 confirmed this and added that management actively discouraged removing headsets rather than encouraging rest breaks.

### Theme 5: Symptoms Are Underreported and Normalized as “Part of the Job”

#### Sub theme 5.1 — No formal reporting pathway used

None of the participants had proactively reported hearing related symptoms through any formal channel (occupational health, HR, or a doctor specifically citing work) before this study. P1’s medical visit was self initiated outside any workplace process, and P3 explicitly confirmed that hearing or ear problems had “never” been raised with anyone before this discussion.

#### Sub theme 5.2 — Peer level awareness without escalation

P3 confirmed noticing similar struggles among coworkers, describing colleagues as “struggling” and “tired,” but reported that this was discussed informally among peers (“that’s how I go”) rather than escalated. This is consistent with a workplace culture that treats these symptoms as an expected cost of the job rather than as an occupational health signal.

### Summary of Themes

Across all three accounts, auditory strain presents as a *cumulative, within shift phenomenon* compounded by unmanaged compensatory volume increases and occurring in a policy vacuum in which no employer guidance on safe headset use exists. Sleep disruption and psychological load appear to interact with auditory symptoms, rather than operating independently of them, particularly in high pressure, target driven roles. Critically, symptom reporting is almost entirely absent from formal channels, which suggests that surveys and self report instruments that rely on participants having already raised concerns with a doctor or employer will substantially undercount prevalence. This supports the study’s premise that a psychological and behavioral early symptom pathway (tinnitus, subjective muffling, perceived ear pressure) may be more sensitive than threshold based audiometry in this population, and it reinforces the case for pairing self report data with objective audiometric testing rather than relying on either alone.

## 2. Discussion

### 2.1 Principal Findings

This study examined self reported auditory dysfunction and its correlates among 46 call center employees in Egypt. Consistent with the study’s first objective, auditory symptoms were highly prevalent: more than three quarters of employees reported mishearing words, difficulty following speech in noise, needing conversations repeated, and heightened sensitivity to loud sounds, and roughly six in ten reported tinnitus. This symptom profile is consistent with early, subclinical auditory strain rather than frank hearing loss, and is broadly in line with what would be expected in an occupational group with prolonged daily headset use and continuous exposure to call related acoustic stimuli.

The composite auditory severity score was approximately normally distributed around a moderate level of symptom burden, with a smaller subset of employees reporting markedly elevated scores (up to 33 out of the composite maximum). This right leaning spread suggests that while most employees experience a low to moderate symptom load, a minority may be experiencing a more concentrated pattern of auditory strain — a group that would benefit most from targeted screening.

### 2.2 Acoustic Shock as the Dominant Risk Factor

Of all the occupational and medical variables tested, a self reported history of acoustic shock — a sudden, unexpectedly loud sound transmitted through the headset — showed by far the strongest and most consistent association with auditory symptom severity, both as a correlate (ρ = .414, p = .004) and as a group comparison (p = .004). This aligns with the broader occupational health literature on call center and telecommunications workers, where acoustic shock incidents (e.g., feedback squeals, line faults, or malicious loud noises from callers) have been identified as a distinct hazard separate from routine background noise exposure (McFerran & Baguley, 2007; Westcott, 2006). The magnitude of this association, in a modest sample, suggests acoustic shock history may be a more sensitive marker of at risk employees than conventional exposure metrics such as years of service or shift pattern — both of which showed negligible correlations with symptom severity in this dataset (ρ = .022 and non significant shift comparison, respectively).

### 2.3 The Auditory–Cognitive Link

The significant correlation between auditory and cognitive symptom severity (ρ = .416, p = .004), driven most strongly by slowed information processing (ρ = .500, p < .001), is consistent with the concept of listening effort: when auditory processing is compromised, cognitive resources are redirected toward decoding speech and monitoring sound, leaving less capacity for other cognitive tasks and manifesting as fatigue, slowed processing, and difficulty concentrating. In a call center setting, where accurate real time listening under noisy conditions is central to job performance, this auditory–cognitive coupling has direct implications for both employee wellbeing and operational performance (e.g., error rates, handle time). The borderline associations for headache and concentration difficulty (p = .053–.061) likely reflect the same underlying mechanism and may reach significance in a larger sample.

### 2.4 Factors That Did Not Emerge as Significant

Contrary to some prior assumptions, shift type was not significantly associated with auditory symptom severity (p = .177), suggesting that when shift is treated as a simple three category variable, it does not capture meaningful variation in auditory risk in this cohort. This may indicate that cumulative or event based exposures (such as acoustic shock or headset volume habits) are more proximally related to symptom severity than the time of day pattern of work itself, or it may reflect limited statistical power to detect a true shift effect. Similarly, years of work and daily/weekly work hour categories showed negligible correlations with severity, which may reflect adaptation over time, survivorship (more severely affected employees may have already left the role), or simply that these coarse categorical exposure measures do not capture cumulative noise dose with sufficient precision.

Headphone/device volume (p = .065) and middle ear disease history (p = .072) both showed trends toward higher severity that did not reach the conventional significance threshold. Given the small size of some subgroups (e.g., only 10 employees reported ear disease), these should be interpreted as hypothesis generating rather than null findings, and would benefit from confirmation in a larger sample. Likewise, the medical history variables that showed no association (cholesterol, hypertension, head trauma) had very small affected subgroups (n = 2–5), so the absence of a significant result more likely reflects insufficient power than a true absence of effect and should not be over interpreted as reassuring.

### 2.5 Limitations

- Cross sectional design: causal or temporal relationships (e.g., whether acoustic shock precedes or follows symptom onset) cannot be established.
- All outcome and exposure measures were self reported; no audiometric testing (pure tone audiometry) was performed to objectively confirm hearing status, so findings reflect perceived rather than measured auditory function.
- Modest overall sample size (N = 46), and several subgroup comparisons involved very small cell sizes (n = 2–10), limiting statistical power to detect true associations for several variables.
- Single center/single country sample, which may limit generalizability to call center workforces with different equipment, noise control policies, or work cultures.
- Recall bias is possible for retrospective exposure items such as acoustic shock history and years of work.
- Composite severity scores, while normally distributed, combine heterogeneous symptoms into a single score, which may obscure symptom specific patterns; nonparametric tests were used throughout for consistency, which is a conservative but not the only defensible analytic choice given the approximately normal distribution of the composite scores.
- Objective audiometric testing was not performed. This decision was informed by prior literature indicating that hearing loss in this occupational population typically does not reach diagnostic clinical thresholds (Pawlaczyk Luszczynska et al., 2018); the study therefore prioritized capturing patient reported auditory and stress symptoms, which may be more sensitive to early or subclinical occupational effects than audiometry alone. However, this means the study cannot confirm or rule out measurable hearing loss in participants, and self reported symptoms are subject to recall and reporting bias. The achieved sample size (n=46) was below the a priori target of 100–120 because reaching the target was not logistically feasible within the data collection period of almost three months; the study is therefore a pilot study, and the small sample limits statistical power and the precision of subgroup comparisons; findings should be interpreted as preliminary. Voluntary (self selected) sampling restricted to three companies via WhatsApp introduces potential selection and non response bias, and many of those reached did not complete the questionnaire, limiting generalizability to the broader call center workforce in Egypt

- Multiple group comparisons were performed at α = .05 without correction for multiplicity, and the sample size did not allow multivariable adjustment for potential confounders (e.g., age, sleep duration, headphone use outside work); the acoustic shock association is therefore best regarded as exploratory.
- There was no comparison group of non call center workers, so the observed symptom frequencies cannot be attributed to call center work alone. The qualitative component comprised only three discussions, and the machine transcribed Arabic–English recordings contained transcription errors, limiting the depth and transferability of the themes

### 2.6 Recommendations

In line with the study’s sixth objective, these findings support several practical, evidence based recommendations for hearing conservation in call center environments:

- Implement or verify acoustic shock (loud noise burst) limiters on all headsets, and establish a formal incident reporting pathway so acoustic shock events are logged and followed up with hearing checks.
- Introduce periodic audiometric screening for employees, prioritizing those who report acoustic shock events or high symptom burden on routine surveys.
- Incorporate brief cognitive symptom screening (e.g., concentration, fatigue) alongside auditory symptom checks, given their observed correlation, to catch early listening effort related strain.
- Provide employee education on safe headset volume levels and encourage regular listening breaks, particularly for employees who report several hours of daily headset use.
- Given the high overall prevalence of early symptoms, consider organization wide awareness campaigns on early warning signs of noise related auditory strain, rather than targeting only high tenure or night shift employees specifically.

### 2.7 Conclusion

In this pilot cross sectional study of 46 call center employees in Greater Cairo, which relied on self reported symptoms, auditory symptoms were common: more than three quarters of participants reported mishearing words, difficulty understanding speech in background noise, needing others to repeat themselves, and sensitivity to loud sounds, and about six in ten reported tinnitus. Symptom severity was most clearly associated with a history of acoustic shock (p = .004) and with concurrent cognitive symptom burden (ρ = .416, p = .004), whereas shift type, years of work, and working hours showed no significant association. Because the subgroups were small and unbalanced (only five day shift employees), a shift effect cannot be excluded, and the borderline trends for headset/device volume (p = .065) and middle ear disease history (p = .072) warrant testing in larger samples. Themes from the interviews were consistent with these findings, describing auditory strain that builds over the shift, compensatory increases in headset volume, disrupted sleep, and no employer guidance or reporting pathway for hearing related symptoms.

These findings should be read as preliminary associations rather than causal effects: the design was cross sectional, all measures were self reported without audiometry, the sample was a self selected volunteer sample smaller than planned (pilot study), and there was no comparison group, so the true prevalence of hearing loss among call center workers cannot be inferred. Even so, they identify acute acoustic events and cumulative listening load, rather than shift schedule or tenure, as the most practical targets for prevention. Acoustic shock limiters with incident reporting, audiometric screening that prioritizes employees with shock events or high symptom burden, safe volume education, and regular listening breaks are recommended, and larger multi site studies pairing symptom questionnaires with audiometry are needed to confirm these results. The response patterns and effect sizes from this pilot study can inform sample size planning for that larger study.

## Data Availability

all data produced in the present whether a questionnaire results and group discussion transcript that been collected through out peroid can be available upon reasonable request to the authors

## Appendix 1. Research Ethics Committee Approval

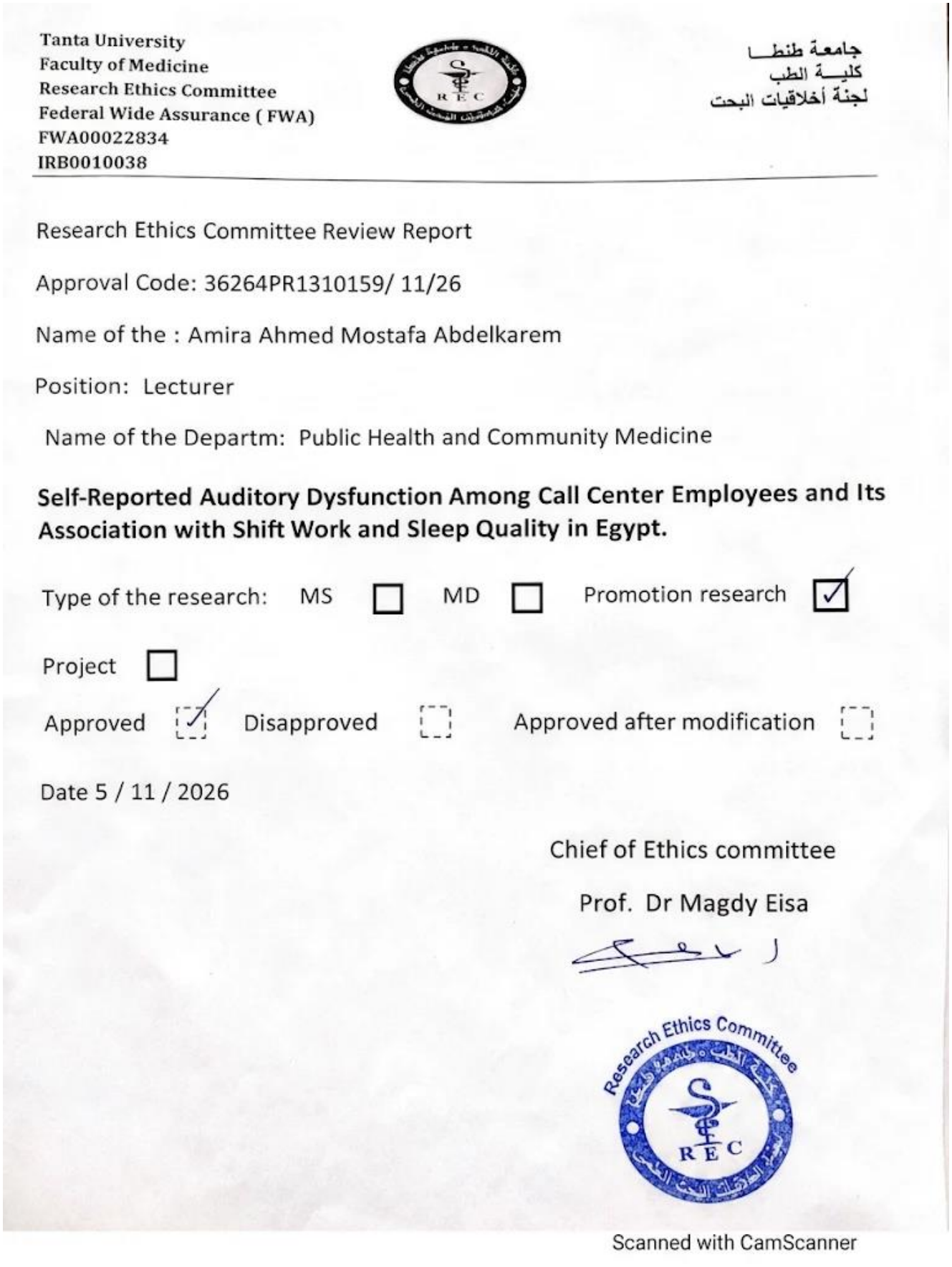

*Research Ethics Committee approval, Faculty of Medicine, Tanta University (5 November 2026)*.

## Appendix 2. Interview Guide

The interviews followed a semi structured guide of open ended questions adapted from questions used in published studies and from standard interview practice. The guide was reviewed by the study supervisor and by the Research Ethics Committee, Faculty of Medicine, Tanta University. A citation beside a question indicates the study from which it was adapted.

### Opening

1. Tell me a bit about your role and how long you have worked here. **Topic 1:** Perceived auditory strain and fatigue
2. Walk me through how your ears feel by the end of a shift compared with the start. What changes do you notice? (Afifi et al., 2020)
3. Have you ever felt that sounds seemed muffled, or that you had to concentrate harder to hear customers later in the day? (Afifi et al., 2020)
4. How does that fatigue affect your work (accuracy, mood, ability to concentrate)?
5. Does it get better on days off, or does it linger? (Venet et al., 2018)
6. Have you noticed any ringing, buzzing, or pressure in your ears during or after calls? **Topic 2:** Sleep disturbance due to night shifts
7. Describe a typical day’s sleep for you. How does it differ on days you work nights versus days off?
8. What is the hardest part of trying to sleep after or before a night shift?
9. How does poor sleep from night shifts show up in your work or home life? (Jeon & Kim, 2022)
10. What have you tried to manage it, and has anything worked?
11. Do you feel more tired, irritable, or forgetful on days after a night shift? **Topic 3:** Anxiety, mental fatigue, coping strategies, and headset use habits
12. During and after your shifts, have you ever experienced symptoms such as nervousness or tension, fatigue, a sense of impending danger, a racing heart, sweating, trembling, difficulty concentrating, or trouble sleeping? Do they recur, and do they continue after the shift?
13. When you start feeling drained or irritated during a shift, what do you actually do to get through it?
14. Tell me about your headset: how you wear it, adjust the volume, or take it off between calls.
15. Does the company give you any guidance on headset volume or breaks, and do you follow it?
16. Have you ever changed your headset habits because of discomfort?
17. Do you talk to coworkers about how to deal with the strain of the job? What kinds of things come up? **Topic 4:** Unreported or subtle symptoms not captured in the questionnaire
18. Is there anything about your hearing or your ears at work that you have never mentioned to anyone (a coworker, a doctor, a survey) until now?
19. Sometimes people notice small things they do not think are “serious enough” to report. Has that happened to you?
20. What stopped you from mentioning it? Did it seem minor, embarrassing, or did you think nothing could be done?
21. Have you noticed anything among coworkers that they do not seem to report either? **Closing**
22. Is there anything about your hearing or health at work that we have not covered and that you think is important?

## Appendix 3. the questionnaire

The questionnaire used to question and study participants working in call centers in Egypt Cairo sampled the already published questionnaire found in already published paper titled Noise exposure and hearing status among call center operators. *Noise and Health*, *20*(96). https://doi.org/10.4103/nah.NAH_11_18 which later been reviewed and validated by the ethical committee in Tanta university public health departments

## References

Afifi, P. O., Abdel Rahman, T. T. T., & Khafagy, A. G. (2020). Auditory fatigue among call center operators with headset. The Egyptian Journal of Otolaryngology, 36(1), Article 41. 10.1186/s4316302000044y

Braun, V., & Clarke, V. (2006). Using thematic analysis in psychology. Qualitative Research in Psychology, 3(2), 77–101. 10.1191/1478088706qp063oa

Chen, K. T., & Su, S. Bin. (2025). A review of occupational noise induced hearing loss: focus on mechanisms and preventive measures. In British Medical Bulletin (Vol. 156, Number 1). 10.1093/bmb/ldaf020

James, S. M., Honn, K. A., Gaddameedhi, S., & Van Dongen, H. P. A. (2017). Shift Work: Disrupted Circadian Rhythms and Sleep Implications for Health and Well Being. Current Sleep Medicine Reports, 3(2), 104–112. 10.1007/S4067501700716

Jeon, B. M., & Kim, S. H. (2022). Associations of extended work, higher workloads and emotional work demands with sleep disturbance among night shift workers. BMC Public Health. 10.1186/s12889022145993

McFerran, D. J., & Baguley, D. M. (2007). Acoustic shock. The Journal of Laryngology & Otology, 121(4), 301–305.

Nelson, D. I., Nelson, R. Y., Concha Barrientos, M., & Fingerhut, M. (2005). The global burden of occupational noise induced hearing loss. American Journal of Industrial Medicine, 48(6), 446–458. 10.1002/ajim.20223

Occupational Safety and Health Administration. (2002). Hearing conservation (OSHA 3074, revised). U.S. Department of Labor.

Pawlaczyk Luszczynska, M., Dudarewicz, A., Zamojska Daniszewska, M., Zaborowski, K., & Rutkowska Kaczmarek, P. (2018). Noise exposure and hearing status among call center operators. Noise and Health, 20(96). 10.4103/nah.NAH_11_18

Toker, M. A. S., & Güler, N. (2022). General mental state and quality of working life of call center employees. Archives of Environmental & Occupational Health, 77(8), 628–635. 10.1080/19338244.2021.1986462

Venet, T., Bey, A., Campo, P., Ducourneau, J., Mifsud, Q., Hoffmann, C., Thomas, A., Mouzé Amady, M., & Parietti Winkler, C. (2018). Auditory fatigue among call dispatchers working with headsets. International Journal of Occupational Medicine and Environmental Health, 31(2), 217–226. 10.13075/ijomeh.1896.01131

Westcott, M. (2006). Acoustic shock injury (ASI). Acta Oto Laryngologica, 126(sup556), 54–58. 10.1080/03655230600895531

